# Childhood Nutrition and Service Delivery Indicators across and between two Epidemics (Ebola and COVID-19) in Sierra Leone: A Descriptive Study using Serial Cross-Sectional Surveys

**DOI:** 10.1101/2025.01.29.25321348

**Authors:** Kadiatu Bangura, Aminata Shamit Koroma, Solade Pyne-Bailey, Manso M Koroma, Sulaiman Lakoh, Stephen Sevalie, Bailah Molleh, Zeleke Abebaw Mekonnen, Adrienne Chan, Sharmistha Mishra, Alhaji U. N’jai

## Abstract

**Introduction:** Sierra Leone is one of the worst affected by hunger and food insecurity, but data to understand the impact of public health emergencies on nutrition indicators is limited. In this study, we sought to describe nutrition service delivery and nutritional health among children under age five before and during the Ebola epidemic, the inter-epidemic period, and during the COVID-19 pandemic (2021) in Sierra Leone.

**Methods:** We conducted a descriptive study using secondary data from five serial cross-sectional surveys conducted using representative sampling as part of programmatic monitoring and evaluation: 2010 (N=14027, before Ebola); 2014 (N=10,975, during Ebola); 2017 and 2019 (N=9059, N=4,870, respectively, inter-pandemic period); 2021 (N=10,165, during COVID-19). We described and compared the prevalence of each of the following indicator at each time-point: shunting, global acute malnutrition, breastfeeding and underweight.

**Results:** The prevalence of stunting was 34.1% before the Ebola epidemic, 28.8% during the Ebola epidemic, 31.3% after the Ebola epidemic, 25.9% before the COVID-19 epidemic, and 26.2% during the COVID-19 epidemic. The prevalence of global acute malnutrition was: 6.9% before the Ebola epidemic, 4.7% during the Ebola epidemic, 5.1% after the Ebola epidemic, 5.0% before the COVID-19 epidemic, and 5.2% during the COVID-19 epidemic. Finally, the report showed that the proportion of breastfeeding for up to 23 months was 84.0% (before the Ebola epidemic), 86.0% (during the Ebola epidemic), 85.0% (after the Ebola epidemic), 61.8% (before the COVID-19 epidemic), and 53.1% (during the COVID-19 epidemic).

**Conclusion:** We found a variable effect of the Ebola epidemic and COVID-19 on nutrition health and nutrition indicators. Findings highlight the importance of continuing to strengthen the implementation of nutrition programs during and after public health emergencies.

## Introduction

Reducing childhood malnutrition is a global health priority, the efforts towards which have been susceptible to health-system disruptions during political, climate, and public health emergencies (1)(2). Globally, an estimated 45 million children under the age of five experienced malnutrition leading to wasting in 2022 (3).

Sierra Leone, as it continues to build its health-system following a decade-long civil war and the largest Ebola virus outbreak in history, remains among the hardest-hit by hunger and food insecurity. In 2023, the country ranked 116^th^ out of 125 on the Global Hunger Index, one in four children under five suffering from stunting (short height for age, reflecting chronic undernutrition) and 5% suffering from global acute malnutrition (4). As a result, the Sierra Leone Ministry of Health identified uninterrupted nutrition services as a key component of the 2019 medium-term national development plan, including preparedness and response to public health emergency (5). In Sierra Leone, a complex set of interconnected factors lead to childhood malnutrition. These factors include, but are not limited to, poverty, low levels of education, poor infant feeding practices, and limited access to potable water (6). Essential community services to optimize nutrition practices include maternal, infant, and young children nutrition awareness and education programme focused on the first 1000 days, from pregnancy to a child’s second birthday. Essential preventative services include monthly nutritional anthropometric screening, micronutrient supplementation and de-worming programmes, and with the combination of treatment services through the Integrated Management of Acute Malnutrition programme (7)(8). This latter program includes community mobilization, targeted supplementary feeding programmes, outpatient therapeutic programmes, and inpatient treatment.

Health-system disruptions can amplify undernutrition. The limited data that exist surrounding health-system disruptions to nutrition services and the health consequences in Sierra Leone suggest concerning patterns (9). A study conducted in a rural district of Sierra Leone found a temporary decline in health-facility screening for malnutrition but community-based screening remained stable during the 2014-2016 Ebola virus outbreak (10). Overall, the rate of diagnoses of acute malnutrition among children doubled after the Ebola outbreak, reflecting a combination of a back-log in diagnoses but also a true increase in the prevalence of acute malnutrition due to disruptions to facility-based services in particular. Thus, when the COVID-19 epidemic hit, the lessons learned from the health-system disruptions caused by the 2014-2016 Ebola virus outbreak, and the interpandemic resilience of health-systems within the country’s post-Ebola recovery efforts were put to the test (11).

In this study, we sought to describe and compare indicators of nutrition service delivery (objective 1), and nutritional health (objective 2) among children under age five across five cross-sectional surveys conducted before and during the Ebola outbreak, interpandemic period, and during the early COVID-19 epidemic in Sierra Leone.

## Methods

### Study design and setting

We conducted a descriptive study using secondary data from five serial cross-sectional surveys in Sierra Leone, and report our findings using the strengthening of the reporting of observational studies in epidemiology guidelines(12). As of 2022, Sierra Leone has a population of 7.5 million of which 1,880,001 are under five children (13).

The Directorate of Food and Nutrition is designed and overseen by the Ministry of Health, and implemented by the Ministry of Health and non-governmental agencies as partners. The programme includes facility- and community-based screening for malnutrition. Nutrition services for children under age five, including screening, outpatient and inpatient treatment are covered under the public system (i.e. there are no user fees).

### Data sources and variables

For this study, we extracted data on indicators from the four National Nutrition Standardized Monitoring and Assessment of Relief and Transition (SMART) surveys (conducted in 2010, 2014, 2017 and 2021) and one Sierra Leone Demographic Health Survey (2019) on June 30, 2023. The data were entered into an Excel database. The five surveys used representative sampling for Food and Nutrition Programme monitoring and evaluation. All the surveys provided the same indicators, measured using the same questionnaire and clinical measurement tools.

First, we extracted study and participant characteristics from each survey: year of survey, proportion of survey participants by age in months, sex and number of children screened for malnutrition in the survey. In the surveys, participants were screened using the same approach as in the national screening programmes.

We address objective 1 (nutritional service delivery indicators) by extracting data on service delivery and uptake: the numerator and denominator of children under five and the proportions of children under five years of age who received vitamin A, children under five years of age dewormed in the six months prior to the survey, infants zero to five months of age who exclusively received breastfeeding, children up to 23 months of age who were breastfeed, children who received timely and appropriate complementary feeding at 6 months, children between 6 and 23 months of age who met the and minimum meal frequency. Furthermore, the nutritional service delivery indicators were measured in the survey through self-report by the guardian of the child. Similarly, to address objective 2 (indicators of nutritional health), we extracted the numerator, denominator (children under five who were screened), and the proportions of stunting, global acute malnutrition, severe acute malnutrition, and underweight.

### Analyses

The five surveys reflect the following periods: pre-Ebola (2010), during Ebola (2014), interpandemic post-Ebola (2017), interpandemic pre-COVID-19 (2019), and during COVID-19 (2021). We reported the descriptive statistics including frequency and percentage for each indicator as outlined above, and characterized patterns in indicators for each objective, by time-period. The analyses were done using Excel and SPSS software.

### Ethics

Scientific Ethics approval was obtained from the Sierra Leone Ethics and Scientific Review Committee, with ethics approval number 014/05/2023. Use of data was approved by the Director of Food and Nutrition and the Chief Medical Officer of the Ministry of Health. The authors did not have access to information that could identify individual participants during or after data collection.

## Results

### Participants and survey characteristics

Overall, 49,096 children under five years of age were surveyed. Of these, 14,027 (28.6%) were recruited in the 2010 survey. The lowest number of children (4,870, 9.9%) were recruited in the 2019 survey. There was no documentation of the gender of children recruited in the 2010 survey, but the other four surveys recruited more female children (17,032, 50.6%) than male children (16,628, 49.4%). The number of districts included in each survey varied because of the administrative structure changes in Sierra Leone over the years (Table 1).

**Table 1:** Participants and Survey characteristics.

| Time Period | Pre-Ebola | During Ebola | Post Ebola | Pre-COVID-19 | During COVID- 19 |
| --- | --- | --- | --- | --- | --- |
| Year of survey | 2010 | 2014 | 2017 | 2019 | 2021 |
| Number of districts | 12 | 12 | 14 | 14 | 16 |
| Number of under five children, screened for malnutrition | 14,027 | 10,975 | 9,059 | 4,870 | 10,165 |
| **Sex** |  |  |  |  |  |
| Male | NA | 5440 | 4486 | 2464 | 4,238 |
| Female | NA | 5535 | 4573 | 2407 | 4,517 |

*NA: Not applicable*

### Pattern of indicators of nutritional service delivery

The proportion of breastfeeding initiation within 1 hour of birth was higher during the Ebola epidemic (54.9%) than before the Ebola epidemic (45%), and showed a gradually increasing trend after the Ebola epidemic (57%), before COVID-19 (75.0%), and during the COVID-19 epidemic (84.9%). The proportion of infants aged 0-5 months who were exclusively breastfed gradually increased from 32.0% before the Ebola epidemic to 58.8% during the Ebola epidemic and 62% after the Ebola epidemic, and then gradually decreased to 54.0% and 52.7% before and during the COVID-19 pandemic, respectively. Vitamin A supplementation was high before the Ebola epidemic (91.1%) and during the Ebola epidemic (96.1%) compared to the inter-epidemic period in 2017 (80.3%) and 2019 (69.0%), but increased to 93.9% during the COVID-19 epidemic. A similar pattern was observed for deworming, which reached its lowest point before the COVID-19 pandemic (64.0%) and increased to 92.9% during the COVID-19 pandemic. In the 2014 survey, indicators for complementary feeding and deworming were not measured. There is a reduction in the proportion of children breastfed up to 23 months from 84.0% pre-Ebola to 53.1% in the COVID-19 period (Table 2).

**Table 2:**
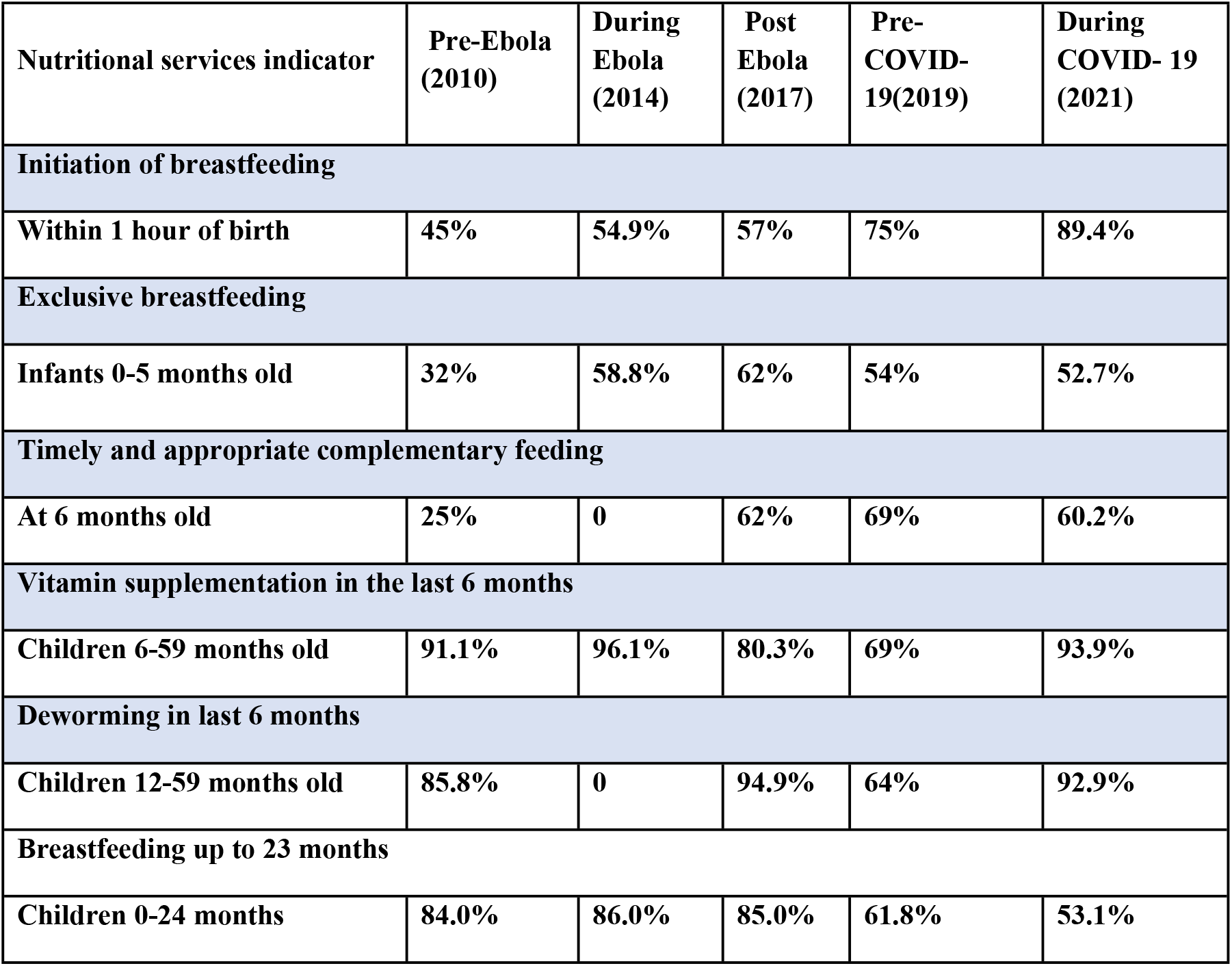
Nutritional services delivery for children under age five years of age across five serial-cross sectional surveys in Sierra Leone.

### Patterns of indicators of nutritional health

The prevalence of stunting fell from 34.1% pre-Ebola to 26.2% during COVID-19. During the intervening years, stunting declined to 28.8% in 2014 (during Ebola), before rising again to 31.3% in 2017 (post-Ebola) and then continuing to decline to 25.9% in 2019 (pre-COVID-19). Similarly, the prevalence of underweight declined from 18.7% in the pre-Ebola period to 11.0% during COVID-19, but rise to 13.6% in the post-Ebola period and 14.0% in the COVID-19 period. In contrast, the prevalence of global acute malnutrition fell during Ebola but has been steadily increasing to stable levels of just over 5% during the interpandemic period into the COVID-19 period ((Table 3).

**Table 3.** Prevalence of Malnutrition and Stunting Among Under-Five children.

| Nutrition Indicators | Pre-Ebola | During Ebola | Post Ebola | Pre-COVID-19 | During COVID- 19 |
| --- | --- | --- | --- | --- | --- |
|  | 2010 | 2014 | 2017 | 2019 | 2021 |
| **Stunting (HAZ <-2)** | **34.1%** | **28.8%** | **31.3%** | **25.9%** | **26.2%** |
| **Global Acute Malnutrition (WHZ <-2)** | **6.9%** | **4.7%** | **5.1%** | **5%** | **5.2%** |
| **Underweight (WAZ <-2)** | **18.7%** | **12.9%** | **13.6%** | **14%** | **11.0%** |

<sup>1</sup> Stunting: children whose height and Age is below -2 standard deviation according to WHO
<sup>2</sup> Global Acute Malnutrition (GAM) are children whose weight when compared to their height is below -2 SD
<sup>3</sup> Underweight are children whose weight when compared to their age according WHO reference is below -2
<sup>4</sup>The periods include: 2010 = pre- Ebola, 2014= during Ebola, 2017= post Ebola, 2019= pre COVID-19 and 2021= during COVID-19. WAZ= Weight-for-age-Z score HAZ=Height-for-age Z-score WHZ=Weight-for-height Z-score

## Discussion

We examined the impact of the 2014/2016 Ebola epidemic and the COVID-19 pandemic on the nutritional indicators and service delivery in children under five in Sierra Leone. The data suggest improvements in most of the indicators of breastfeeding and complementary feeding increased significantly from the pre-Ebola period to the post-Ebola period and during COVID-19. We also found that the prevalence of stunting decreased from 34.1% pre-Ebola to 26.2% during the COVID-19 pandemic, but increased during the inter-epidemic period (post-Ebola and pre-COVID-19). The prevalence of global acute malnutrition increased from 4.7% during the Ebola epidemic to 5.2% during the COVID-19 pandemic. Finally, there is a reduction in the proportion of children breastfed up to 23 months from 84.0% pre-Ebola to 53.1% in the COVID-19 period.

There is a well-established relationship between malnutrition and public health emergencies, particularly Ebola and COVID-19 (10) (14). Disruptions in supply and agricultural value chains, food supplements and other essential services, and socioeconomic shocks caused by pandemics or epidemics are among the factors that contribute to malnutrition in children under five during public health emergencies (14). Sierra Leone’s persistent poverty and hunger levels with high rural-urban, age and gender disparities in these indices exacerbate malnutrition in public health emergencies (4, 6).

Nutrition surveys conducted before the Ebola outbreak had the largest number of participants compared to other nutrition surveys. While this may be attributable to the methodologies used in different surveys, it may also be due to community perceptions, misinformation, fear, and lack of trust in the health system during and after the epidemic.

Our study showed improvements in many indicators of nutrition service delivery, such as initiation of breastfeeding within 1 hour of birth, exclusive breastfeeding for infants 0-5 months, and vitamin A supplementation. These changes underscore the improvement in the nutrition programme quality and expansion of nutrition services such as scaling up outpatient treatment programmes, mothers-to-mother support groups on infant and young children feeding practices and development and dissemination of nutrition policies (16). For example, large-scale door-to-door vitamin A distribution campaigns may explain the high rate of vitamin A supplementation among children aged 6–59 months during the Ebola epidemic. All of these interventions, as well as lessons learned from the Ebola outbreak, may have also helped to increase the resilience of the system to screen, manage and prevent nutritional disorders (15).

Although the COVID-19 pandemic is expected to increase the risk of various forms of malnutrition, the prevalence of stunting and underweight among children under five years of age was lower during the COVID-19 pandemic than during the Ebola epidemic (17). These dynamics may be explained by differences in the severity of the two diseases and their associated impacts (18, 19). Due to the higher mortality rate of Ebola epidemic, stringent restrictions and consequently essential service disruptions were more stringent than those of the COVID-19 pandemic (20).

Unlike stunting and underweight, the prevalence of global acute malnutrition was higher during COVID-19 than in Ebola. Global acute malnutrition is a measure of a population’s nutritional status and an indicator of an acute food shortage (21). The lower prevalence of global acute malnutrition during the Ebola epidemic compared with the COVID-19 pandemic can be explained by the higher external support during the Ebola epidemic, which mitigated the nutritional impact.

The reduction in the proportion of children breastfed up to 23 months from 84.0% pre-Ebola period to 53.1% in the COVID-19 period may be due to the change in the global and national recommendation for continued breastfeeding from 12 months to 23 months as health workers and mothers needed time to adapt to the new guidelines (22). We recommend that the Ministry of Health focus attention on this indicator by providing job aids, mentorship and other learning opportunities for health service providers.

Our study has limitations inherent to retrospective studies. The use of secondary data limited the study to a descriptive rather than a mixed-method design to capture caregivers’ perspectives. Comparison of data across regions and districts is not possible due to gaps and inconsistencies in some of the data collected. We used aggregated data which makes it impossible to do regression analysis to understand the determinants of poor nutrition indicators during COVID-19. Nonetheless, our study has provided data on the impact of COVID-19 on the nutrition services and indicators in Sierra Leone.

## Conclusion

Overall, we found that most nutrition indicators, such as stunting and underweight among children under five, worsened after the Ebola outbreak but improved during the COVID-19 pandemic. We recommend that low-income countries continue to strengthen the implementation of nutrition programs during and after public health emergencies.

## Data Availability

The data (https://doi.org/10.5061/dryad.4tmpg4fn0) analyzed for this study is available online from: https://datadryad.org/stash/dashboard

## Acknowledgements

We acknowledge the staff of the Directorate of Food and Nutrition in the Ministry of Health for the support in the conduct this study. We thank Kristy Yiu (Unity Health Toronto) for her early support in coordination of the project. Finally, we thank the staff of Sustainable Health System for their support. SM is supported by a Tier 2 Canada Research Chair in Mathematical Modeling and Program Science.

## Funding

This study was funded supported by the Canadian Institute of Health Research (grant number: CIHR: WI1-179883).

## Conflict of interest

The authors declare no conflict of interest.

## Author’s contribution

Conceptualization and study design: ASK, KB and AUN

Data curation and validation: BM, KB

Methodology and formal analysis: AUN, BM, MMK, ZBM

Funding acquisition, resources, supervision: SS, SL, SM, ACK

Writing – original draft preparation: KB, ASK, MMK, SP, AUN

Writing – review & editing: KB, SL, AC, SM, SS

## Notes

### Competing Interest Statement

The authors have declared no competing interest.

### Author Declarations

Scientific Ethics approval was obtained from the Sierra Leone Ethics and Scientific Review Committee, with ethics approval number 014/05/2023.

